# Diurnal variation in SARS-CoV-2 PCR test results: Test accuracy may vary by time of day

**DOI:** 10.1101/2021.03.12.21253015

**Authors:** Candace D. McNaughton, Nicholas M. Adams, Carl Hirschie Johnson, Michael J. Ward, Thomas A. Lasko

## Abstract

False negative tests for SARS-CoV-2 are common and have important public health and medical implications. We tested the hypothesis that the proportion of positive SARS-CoV-2 real-time polymerase chain reaction (RT-PCR) tests varied by time of day, suggesting variation in viral shedding by time of day. Among 30,000 clinical tests performed among symptomatic and asymptomatic patients in the Vanderbilt Affiliated Healthcare Network from March-June 2020, we found evidence for diurnal variation in the proportion of positive SARS-CoV-2 tests, with a peak around 2pm in the afternoon and 2-fold variation over the day. Variation was most pronounced in outpatient and inpatient testing locations. These findings have important implications for public health testing and vaccination strategies.

## INTRODUCTION

Optimal diagnostic testing strategies are a key component in effective measures to control spread of disease (1). False negative tests may occur due to sample collection techniques, test sensitivity, and time point in the clinical course (2–4). Anecdotal variation in symptoms of COVID-19 by time of day suggests viral shedding may be cyclical, but previous work has not explicitly examined this. We therefore sought to test the hypothesis that the proportion of positive SARS-CoV-2 RT-PCR tests varied by time of day.

## METHODS

We extracted results of all SARS-CoV-2 RT-PCR tests from the electronic medical record of the Vanderbilt Health Affiliated Network, which is a regional health system in central Tennessee in the United States with a catchment area of 65,000 square miles serving >300,000 patients. Nasopharyngeal swab samples were collected by clinical staff in outpatient, inpatient, and emergency department locations beginning in early March through late June 2020 (exact dates are unavailable due to data de-identification). In accordance with clinical guidelines, approximately the first 18,000 tests (about 40%) were performed for only symptomatic patients; in late April 2020, screening of all asymptomatic hospitalized patients was initiated as well. Test results, time stamp and clinical location, as well as patient age and sex, were obtained from the electronic medical record; there were 93 (0.2%) indeterminate RT-PCR results, which were excluded. The institutional Human Subjects Research Protection Program approved this study as non-human subjects research.

RT-PCR was conducted according to the FDA Emergency Use Authorized (EUA) CDC 2019-nCoV Real-Time RT-PCR Diagnostic Panel in a CLIA-accredited laboratory. Primer and probe sets for N1, N2, and N3 (targeting three regions of SARS-CoV-2 nucleocapsid gene) and RP (an internal human RNase P control) obtained from Integrated DNA Technologies and TaqPath 1-Step qRT-PCR enzyme mixture purchased from Life Technologies were used to perform RT-PCR. Initially, all four primer and probe targets were used. After March 16, 2020, the N3 target was removed from the assay in accordance with an update to the CDC EUA testing protocol. Thermocycling was performed with FAM-signal normalized against ROX according to the following cycling conditions: 25 ° C for 2 min, 50 ° C for 15 min, 95 ° C for 2 min, then 45 cycles of 95 ° C for 3 s and 55 ° C for 30 s. A positive test result was reported for specimens that yielded positive signal for all N-gene targets (C_q_ < 40). A single batch of approximately 1,400 samples were sent to an external lab near the end of March because of high test volumes, and similar RT-PCR methods were used.

The fraction of positive tests was examined by time of day using kernel density estimation with a 1-hour kernel bandwidth, with 95% confidence intervals computed using 10,000X bootstrap resampling, and a sinewave was fit by nonlinear least squares to the median curve (pandas v1.0.3 scikit-learn, scipy.optimize). Testing sites (inpatient, outpatient, and emergency department) were also examined separately.

## RESULTS

We assessed 31,094 RT-PCR test results for SARS-CoV-2 collected by nasopharyngeal swabs from 28,101 patients ≥ 18 years old across 127 testing sites over ∼12 weeks within a regional healthcare system in the United States. Of these, 2,438 (7.8%) were positive for SARS-CoV-2. Between 16 and 1,232 tests per day were performed; the fraction positive varied between 0 and 18%, and 27,561 (88.6%) tests were performed between 8 am and 8 pm. Demographics were similar among outpatient sites (Table); patients in the emergency department and hospital were older than outpatients, and 65.9% of tests were for White patients, compared to the local community prevalence of ∼80%. A 24-hour sinusoidal variation in the fraction of positive tests was observed, with a peak in the fitted sinusoid at 1:49 pm, and 2.2-fold difference between the fitted peak and trough (Figure 1a). Less temporal variation was noted for tests performed in the emergency department.

**Table :** Characteristics of administered SARS-CoV-2 RT-PCRT nasopharyngeal tests

|  | All<sup>a</sup> | Outpatient | Inpatient | Emergency Department |
| --- | --- | --- | --- | --- |
| Number of testing sites | 127 | 60 | 55 | 3 |
| Number of tests | 31,094 | 23,480 | 5,732 | 1,721 |
| Female, N (%) | 17,705 (56.9%) | 13,819 (58.9%) | 2,894 (50.5%) | 899 (52.2%) |
| Age (years) median [IQR] | 43.3 [30.9, 58.6] | 41.4 [30.2, 56.0] | 56.2 [37.2, 68.9] | 37.1 [28.0, 50.7] |
| Race, N (%) |  |  |  |  |
| Asian | 613 (2.0%) | 479 (2.0%) | 83 (1.4%) | 50 (2.9%) |
| Black | 3,660 (11.8%) | 2,106 (9.0%) | 1,111 (19.4 %) | 428 (24.9%) |
| Native American | 53 (0.2%) | 40 (0.2%) | 7 (0.1%) | 5 (0.3%) |
| Multiple | 152 (0.5%) | 93 (0.4 %) | 45 (0.8%) | 13 (0.8%) |
| White | 20,495 (65.9%) | 15,086 (64.3 %) | 4,212 (73.5%) | 1,069 (62.1%) |
| Other | 558 (1.8%) | 276 (1.2%) | 186 (3.2%) | 95 (5.5%) |
| Unknown | 5,563 (17.9%) | 5,400 (23.0%) | 88 (1.5%) | 61 (3.5%) |
| SARS-CoV-2 detected by RT-PCR | 2,438 (7.8%) | 1,992 (8.4%) | 297 (5.1%) | 147 (8.5%) |

<sup>a</sup> Small site categories omitted, thus subgroups do not sum to 'All'.
Abbreviations: IQR, interquartile interval; RT-PCR, real-time polymerase chain reaction

**Figure :**
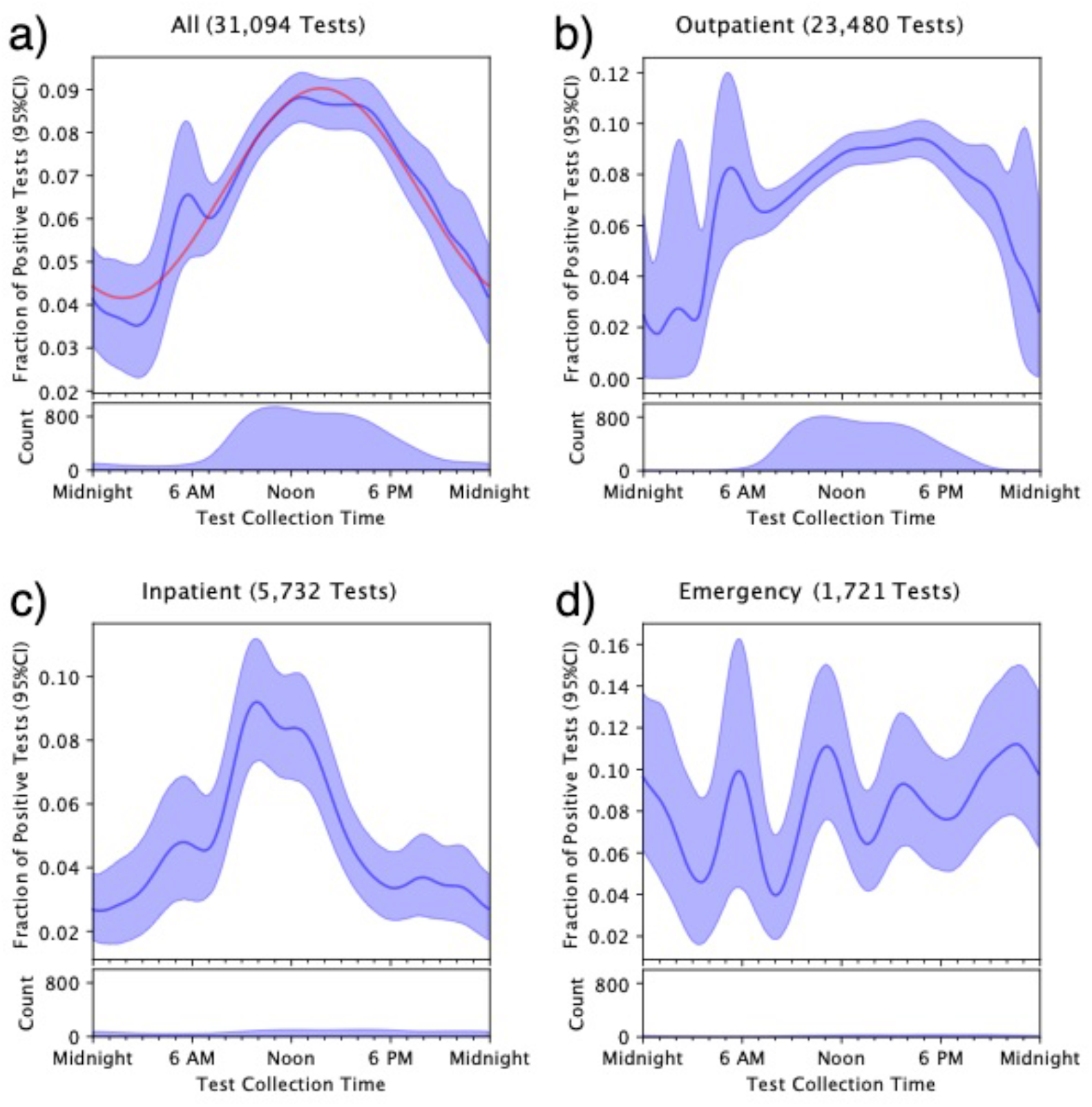
Fraction of positive tests by time of day. The dark blue line is the median curve after bootstrap resampling, with 95% confidence intervals shaded. The red curve in panel (a) is the best fit sinewave, which peaks at 1:49 pm. Note: scale for number of tests performed differs by panel.

The strong diurnal sinusoid pattern in the fraction of positive SARS-CoV-2 tests suggests a cyclic pattern in viral shedding, with a peak in the early afternoon. This observation aligns with previous findings for other viruses such as influenza, herpes, and dengue, where interactions with the immune system lead to diurnal variation in viral shedding and symptoms (5–7), and have been shown to influence vaccine effectiveness (8, 9).

Although we cannot exclude the risk of potential confounding by indication, if asymptomatic patients were preferentially tested during certain hours, the fraction of positive tests would likely be diluted during those times (Figure 1c); of note, asymptomatic hospitalized patients were tested when the decision to admit was made, not at pre-determined testing times. Alternatively, if emergency department patients were more likely to be symptomatic or farther into the disease course, we would expect them to be more likely to have continuously detectible viral RNA in the nasopharynx (4) and bias our results towards the null (Figure 1d).

Potential diurnal variation in SARS-CoV-2 warrants further investigation because, if confirmed, it has important implications for public health and diagnostic strategies. First, a 2-fold variation in test sensitivity could be used to optimize test collection time, result interpretation, and patient counselling. Additionally, a shedding cycle that peaks in the afternoon may play a role in community and hospital spread if it occurs when patients are more likely to interact with others or seek medical care (10). Such temporal considerations may be leveraged to maximize the effectiveness of non-pharmaceutical intervention strategies to contain further viral spread and even vaccination strategies.

## Data Availability

Data not available publicly due to legal restrictions.

